# Severity Prediction for COVID-19 Patients via Recurrent Neural Networks

**DOI:** 10.1101/2020.08.28.20184200

**Authors:** Junghwan Lee, Casey Ta, Jae Hyun Kim, Cong Liu, Chunhua Weng

**Affiliations:** Department of Biomedical Informatics, Columbia University, New York, N.Y.

## Abstract

The novel coronavirus disease-2019 (COVID-19) pandemic has threatened the health of tens of millions of people worldwide and imposed heavy burden on global healthcare systems. In this paper, we propose a model to predict whether a patient infected with COVID-19 will develop severe outcomes based only on the patient’s historical electronic health records (EHR) prior to hospital admission using recurrent neural networks. The model predicts risk score that represents the probability for a patient to progress into severe status (mechanical ventilation, tracheostomy, or death) after being infected with COVID-19. The model achieved 0.846 area under the receiver operating characteristic curve in predicting patients’ outcomes averaged over 5-fold cross validation. While many of the existing models use features obtained after diagnosis of COVID-19, our proposed model only utilizes a patient’s historical EHR to enable proactive risk management at the time of hospital admission.

## INTRODUCTION

The novel coronavirus disease-2019 (COVID-19) has threatened the health of tens of millions of people over the world and imposed heavy burden on global healthcare systems. To fight against the pandemic and mitigate the burden, numerous efforts have been made by scientists to develop risk prediction models for COVID-19 patients. Prognostic models, among the important risk prediction models, has been developed to predict risks of mortality^1-3^ and progression to severe status^4-6^ for COVID-19 patients. Commonly used predictors for those COVID-19 prognostic models include comorbidities, age, sex, lab test results (e.g., lymphocyte count, C reactive protein, and creatinine), and radiologic imaging features^7^. The existing models, however, spanning from Cox proportional hazards models to state-of-the-art machine learning and deep learning models, heavily rely on features obtained after hospital admission or diagnosis of COVID-19 for post-diagnosis prognosis^7^.

Recurrent neural networks (RNN) have been widely used in modeling sequential phenomena such as speech and language due to its strengths of capturing hidden relationships between the sequential data^8^. There have been several studies in the healthcare domain that used RNN to predict future medical events or the risk of certain diseases, leveraging the sequential nature of electronic health records (EHR). For example, Lipton et al.^9^ and Choi et al.^10^ both used RNN for predicting future medical events based on the historical EHR data. Choi et al.^11^ published related work using RNN for predicting the risk of heart failure based on the patient’s historical EHR data.

In this study, we applied RNN on a patient’s historical EHR data to predict the patient’s risk of developing severe outcomes from COVID-19, including mechanical ventilation, tracheostomy, or death. The prediction represents the probability for a patient to progress into a severe status after being infected with COVID-19. One major advantage of this method is that the model does not require any data after the diagnosis of COVID-19 (e.g., lab test results and vital signs), so that it can predict the risk of developing severe outcomes from COVID-19 for a patient before or at the time of hospital admission. This advantage allows proactive risk management by the clinical care team and resource allocation in advance, which can be critical for health policy makers and hospital administrators.

## METHODS

### COVID-19 Cohort Description

New York City has been one of the epicenters of the COVID-19 pandemic. NewYork Presbyterian Hospital/Columbia University Irving Medical Center (NYP/CUIMC) has treated a large cohort of COVID-19 patients since the onset of the pandemic. For this work, we obtained all EHR data for the patients infected with COVID-19 updated until May 31, 2020 from NYP/CUIMC’s Observational Medical Outcomes Partnership (OMOP) database, which contains 30 years’ worth of comprehensive EHR data for about 6.5 million patients. This work received institutional review board approval (AAAR3954) with a waiver for informed consent.

The COVID-19 cohort was identified as patients 18 years or older who were hospitalized and tested positive for SARS-CoV-2 within 21 days before or during their hospitalization. The patients must have at least one visit record prior to March 1, 2020 and with at least one condition (i.e. diagnosis) concept. We obtained all condition concepts in historical inpatient and outpatient visits prior to the hospital admission due to infection of COVID-19 for the identified patients in the cohort in temporal order. In total, 5,774 unique condition concepts were identified from all patients in the cohort. Demographic information (i.e. sex and age at the most recent hospital admission) of the patients were also obtained. Characteristics of the COVID-19 cohort are shown in **Table 1**. We classified patients in the COVID-19 cohort into two groups: severe vs. moderate. Severe patients were identified as the patients who had at least one of the following outcomes during hospitalization: mechanical ventilation, tracheostomy, or death; these events correspond to a severity score of ≥ 6 in the World Health Organization ordinal scale for clinical improvement^12^. Moderate patients refer to the patients who were either discharged without developing severe outcomes during hospitalization or were still hospitalized but without any signal of the severe outcomes.

**Table 1.** Characteristics of the COVID-19 cohort. Senior patients: aged ≥ 65 at the most recent hospital admission. SD: standard deviation.

|  | Severe patients | Moderate patients |
| --- | --- | --- |
| Total # of patients | 546 | 1,828 |
| # of patients with either mechanical ventilation or tracheostomy | 15 | - |
| # of death | 531 | - |
| # of senior patients (%) | 455 (83.3%) | 878 (48.0%) |
| male patients (%) | 322 (59.0%) | 891 (48.7%) |
| Avg. age of the patients (SD) | 76.79 (12.92) | 61.34 (18.29) |
| Median # of visits per patient (25 percentile, 75 percentile) | 16.0 (4.0, 46.0) | 12.0 (3.0, 38.0) |

### Problem Definition

For notation, we denote vectors with italic bold lower-case (e.g. ***h*_1_, *x*_1_**), matrices with italic bold upper-case (e.g. ***W***_***FC***_), and scalars with italic lower-case (e.g. *ŷ*). For notational convenience, we assume that the input for the model is a single patient.

For each patient in the cohort, all historical inpatient and outpatient visits were extracted in the form of multi-hot encoded vector ***x***_***i***_ for *i* = 1,, *p*, where *p* is the number of total visit that the patient made before the hospital admission due to COVID-19. Inpatient visits included emergency room visits or hospitalizations via emergency room. The multi-hot encoded vector ***x***_***i***_ ∈ [0, 1]^*k*^ represents the *i*-th visit of the patient, where *k* denotes the number of unique medical concepts observed in the cohort. 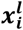 is 1 if the *l*-th medical concept was observed in the patient’s *i*-th visit and 0 otherwise. Our goal is to predict a patient’s risk of developing severe outcomes based on the patient’s historical EHR data. The predicted risk score ranges between 0 and 1 and represents the estimated probability for the patient to progress into a severe outcome from COVID-19.

### Model Architecture

The proposed RNN model to predict the risk score is depicted in **Figure 1**. At each timestamp i, the model receives a patient’s visit x and the previous hidden state *h*_*i*−1_ as input and outputs hidden state *h*_*i*_ for *i* = 1,, *p*, where *p* is the number of total visit that the patient made. We used Gated Recurrent Units^13^ (GRU) for the RNN model in this work. Although Long Short Term Memory^14^ (LSTM) is the most widely used RNN cell among all other RNN variants and generally outperforms GRU on large datasets^15,16^, GRU show comparable or better performance on tasks with relatively small datasets with fewer parameters^17^. Preliminaries of GRU are available in the supplementary material.

**Figure 1.**
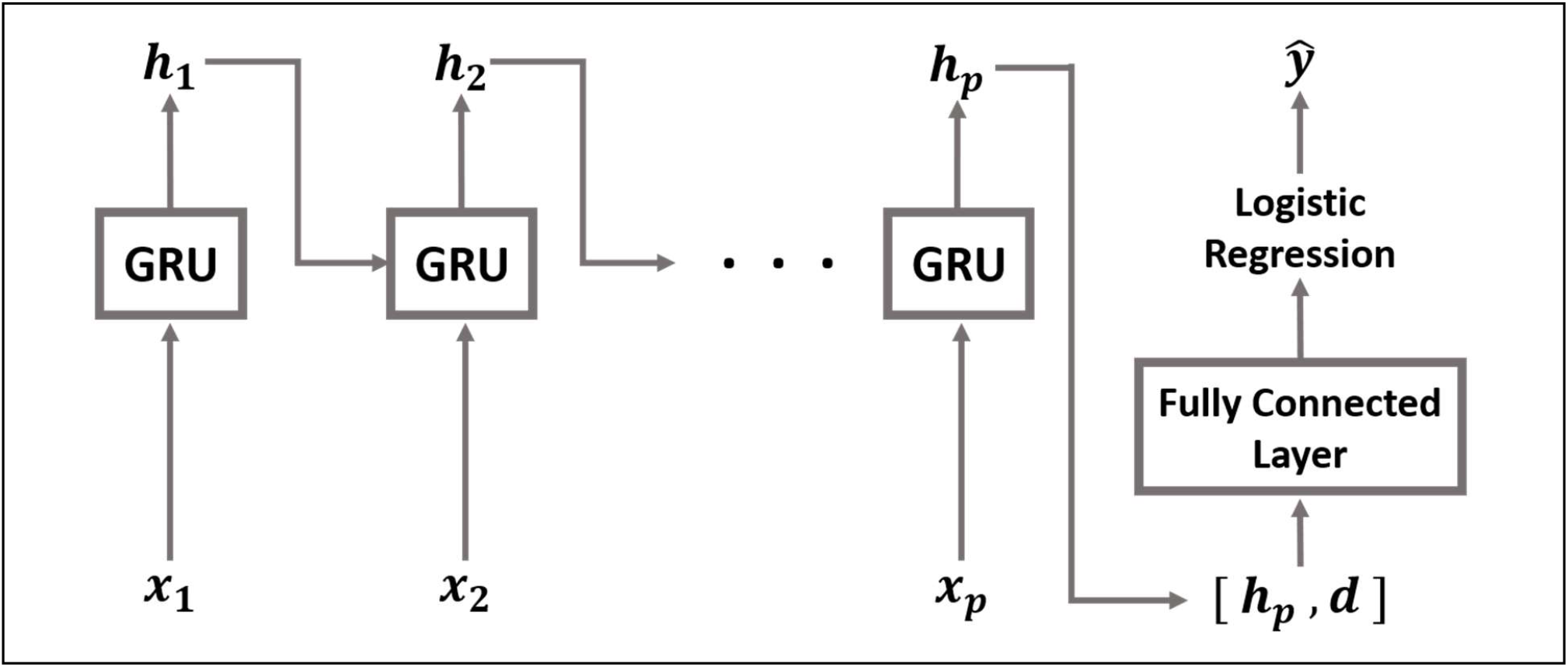
The architecture of the proposed recurrent neural network model. GRU: Gated Recurrent Unit.

For efficient training of the model, we used an embedding layer that transforms the multi-hot encoded input ***x***_***i***_ into a low-dimensional embedding (described below). The hidden state at the last timestamp is concatenated with the patient’s demographic information vector and subsequently fed into a fully connected layer with hyperbolic tangent activation. Finally, an output layer that contains a single neuron with sigmoid activation (i.e. logistic regression layer) is applied over the output of the fully connected layer to generate the risk score of the patient as defined in Eq(1) and Eq(2):

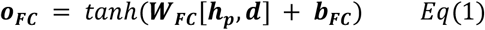

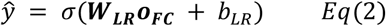

where ***W***_***FC***_, ***W***_***LR***_, ***b***_***FC***_, *b*_*LR*_, *o*_*FC*_, *h*_*p*_, *d* and *ŷ* denote the weight matrix of the fully connected layer, weight matrix of the logistic regression layer, bias of the fully connected layer, bias of the logistic regression layer, the output vector of the fully connected layer, the hidden state at the last timestamp *p*, demographic information vector, and the predicted risk score of the patient respectively. [·,·] denotes vector concatenation, *tanh*() denotes the hyperbolic tangent activation function, and *σ*() denotes the sigmoid activation function. A patient’s demographic information vector is a simple concatenation of one-hot encoded sex (i.e. [1, 0] for male and [0, 1] for female) and min-max normalized age of the patient.

The true label *y* for each patient was determined based on outcome status of the patient as observed in the CUIMC database: we assigned 1 for severe patients and 0 for moderate patients. Since severe and moderate cases were imbalanced in the dataset, we used weighted cross entropy loss, defined as Eq(3):

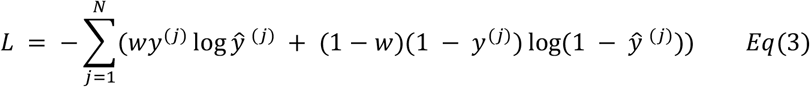

where *y*^(*j*)^, *ŷ*^(*j*)^, *N*, and *w* are the true label for the *j-*th patient, the predicted risk score for the *j-*th patient, the total number of patients in the batch, and weight for the cross entropy. We used 0.75 for the weight of the cross entropy considering the ratio of the severe and moderate patients in the cohort to provide more weight on accurately predicting severe cases (i.e. more focus on sensitivity).

## RESULTS

### Experiment Setup

To evaluate the performance of the RNN model, we compared the average area under the receiver operating characteristic curve (AUC) based on 5-fold cross validation with two other baselines – logistic regression and multilayer perceptron (MLP). The entire dataset was divided into 5 chunks: 3, 1, and 1 chunk(s) were allocated to the training set, validation set, and test set respectively (i.e. 60% training, 20% validation, 20% test split). Different combinations of chunks were allocated to the training set, validation set, and test set at every fold, thus the model was trained, validated, and tested on different datasets at every fold. All models were trained with a maximum of 50 epochs at every fold and the model achieved the highest AUC on the validation set was finally used for test set evaluation. We reported the average and standard error of AUCs of all 5 folds based on the test set.

We used an embedding layer to transform the multi-hot encoded input ***x***_***i***_ into a low-dimensional embedding. We experimented with two different initializations of the embedding layer in the RNN model: (1) the embedding layer initialized with a random normal distribution; (2) the embedding layer initialized with pre-trained embedding. Random normal distribution with mean 0 and standard deviation 0.01 was chosen for initialization since it showed better performance than many other baselines in word embedding tasks^18^. We pre-trained an embedding using GloVe^19^ on the co-occurrence matrix obtained from the cohort for 100 epochs. The pre-trained embedding captures the relationships between the medical concepts since GloVe utilizes the global co-occurrence matrix of concepts for its training, where the co-occurrence matrix is calculated based on the concept co-occurrence in every patients’ visit. In the literature, the dimensionality of the embedding is generally set between 100-500 for medical concept vocabularies with sizes from a few hundred to tens of thousands of concepts^11,20^, therefore we set the dimensionality of the pre-trained embedding and randomly initialized embedding to 128. The embedding layer was fine-tuned jointly with the prediction task of the model.

Since mini-batch training shows good generalization performance when the size of the data is relatively small^21^, we used a small batch of size 2 in the training. We also empirically found that prediction performance of the model decreased with larger batch size. To prevent the model from overfitting, *L*_2_ weight decay with regularization coefficient of 0.001 was applied to weights of the fully connected layer in the RNN model. We tried dropout^22^ to non-recurrent connection of the RNN model and found that dropout did not improve the performance of the model, therefore we did not use dropout.

## Baselines

### Logistic regression

A simple logistic regression model was used for the first baseline with three different types of input: aggregated multi-hot encoded vector, aggregated embedding, and aggregated pre-trained embedding. For each patient, aggregated multi-hot encoded vector is summation of input *x*_*i*_ at all timestamps, after which is clipped with maximum value to 1. Aggregated embedding and aggregated pre-trained embedding were generated by passing the aggregated multi-hot encoded vector through randomly initialized embedding layer or pre-trained embedding layer respectively. Those two embedding layers were initialized using the same scheme as the RNN model. Aggregation of the input can be understood as the summation for each concept observed across visits in a patient’s history. All aggregated inputs were normalized to zero mean and unit variance for numeric stability during training. *L*_2_ weight decay with regularization coefficient of 0.001 was applied to weights in the model to reduce overfitting.

### Multilayer perceptron

Multilayer perceptron (MLP) with a single hidden layer was used for another baseline. A fully connected layer with hyperbolic tangent activation was used for the hidden layer and the output layer contains a single neuron with sigmoid activation. The three different types of inputs (aggregated multi-hot encoded vector, aggregated embedding, and aggregated pre-trained embedding) were used with the same settings as described above. The number of hidden units in the hidden layer was set to 128. *L*_2_ weight decay with regularization coefficient of 0.001 was applied to weights in the model to reduce overfitting.

### Implementation Details

We used Tensorflow 2.0.0^23^ to implement the RNN model and all baselines. Adam^24^ was used for optimization in training for all models. A machine equipped with 2 × Intel Xeon Silver 4110 CPUs and 192GB RAM was used. Hyperparameters and some important details of training are provided in the supplementary material. The source codes to implement all models are publicly available at https://github.com/Jayaos/rnn-covid.

### Prediction Performance of the Risk Score

We calculated the average AUC of 5-fold cross validation to evaluate the prediction performance of the risk score generated by the models (**Figure 2**). Overall, the RNN model with pre-trained embedding achieved the highest average AUC (0.846). The RNN model also showed higher average AUC than the baselines when comparing the same embedding layer initialization schemes.

**Figure 2.**
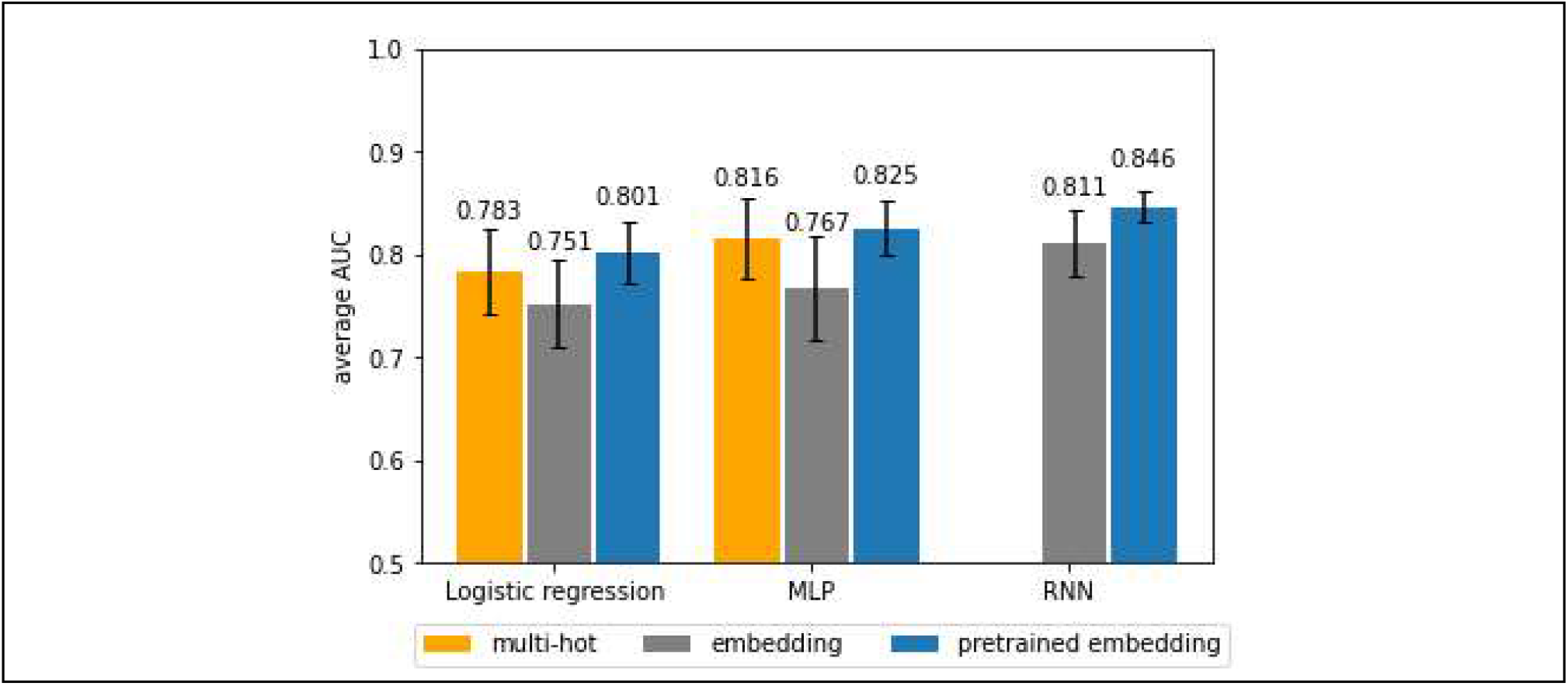
Average 5-fold cross validation AUC of all models. The values of average AUC and standard error of AUC for each model are provided in the supplementary material.

### Prediction Time

For the RNN model with pre-trained embedding, which showed the best performance, approximately 0.017 seconds were required to make a prediction for a single patient. We measured the time by averaging the time that the model took to make predictions on the entire test set using the same machine as described in the Implementation Details section.

### Analysis of the Risk Score

We analyzed the risk score generated by the RNN model with basic characteristics of the patients to understand how the risk score is affected by patient characteristics. Age and historical visit count were selected as baselines since they were expected to serve as proxies of a patient’s general health status. We used the best performing model among the five models (RNN with pre-trained embedding) in 5-fold cross validation to obtain the risk score of the patients in the test set of the corresponding fold. **Figure 3** shows the scatterplot between the risk score and (**a**) outcome status (**b**) age (min-max normalized age), and (**c**) historical visit count of the patients. The regression coefficient were +0.136 (p < 0.01) in (**a**), +0.376 (p < 0.01) in (**b**), and +0.0002 (p < 0.01) in (**c**). **Figure 4** shows the ROC curve of the risk score, age, and historical visit count in predicting the outcome status of the patients.

**Figure 3.**
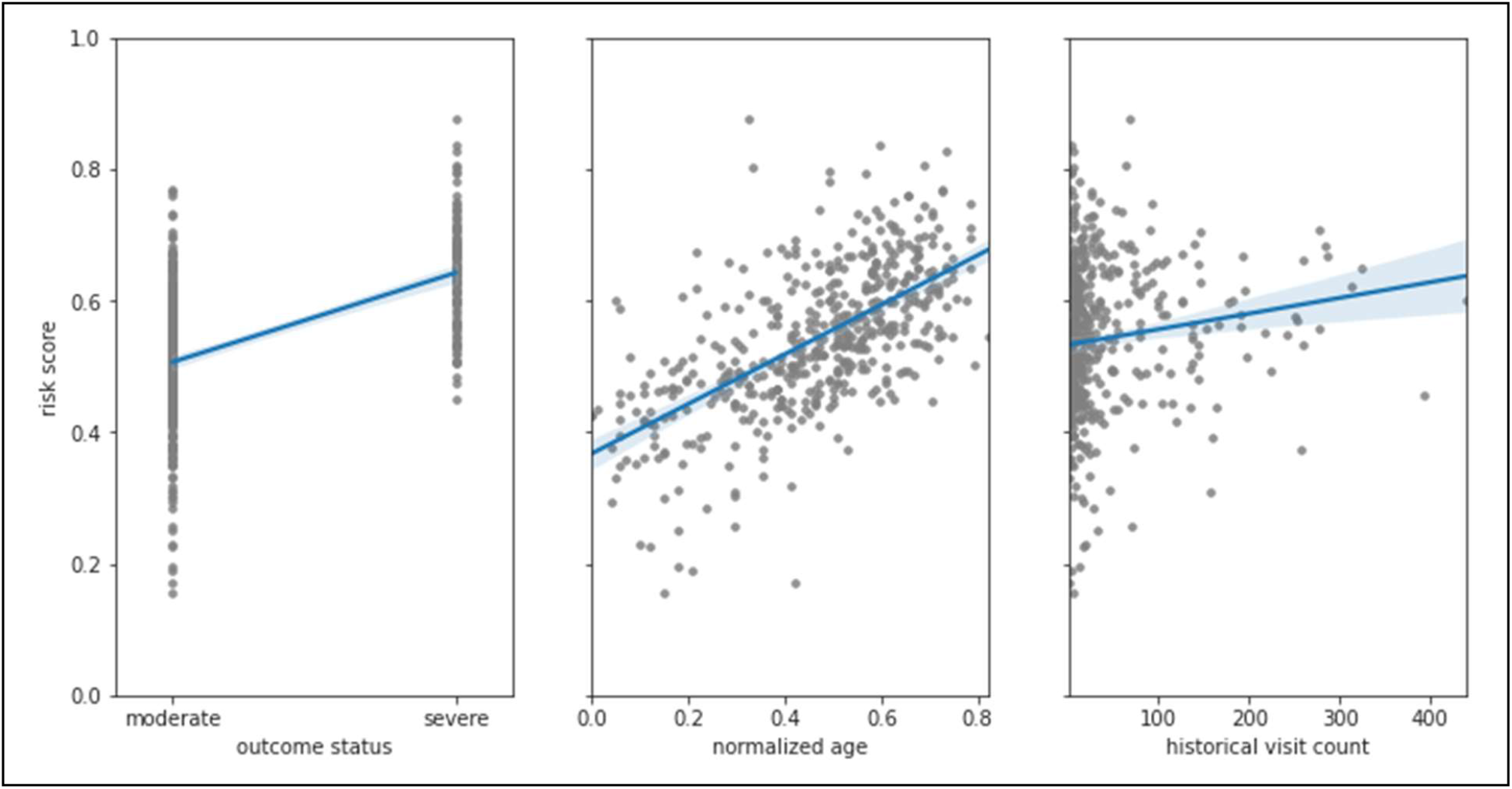
Scatterplot of (**a**) the outcome status and the risk score, (**b**) normalized age and the risk score, and (**c**) historical visit count and the risk score with the regression line. The gray-colored dots represent patients and shaded region around the regression line represents confidence interval.

**Figure 4.**
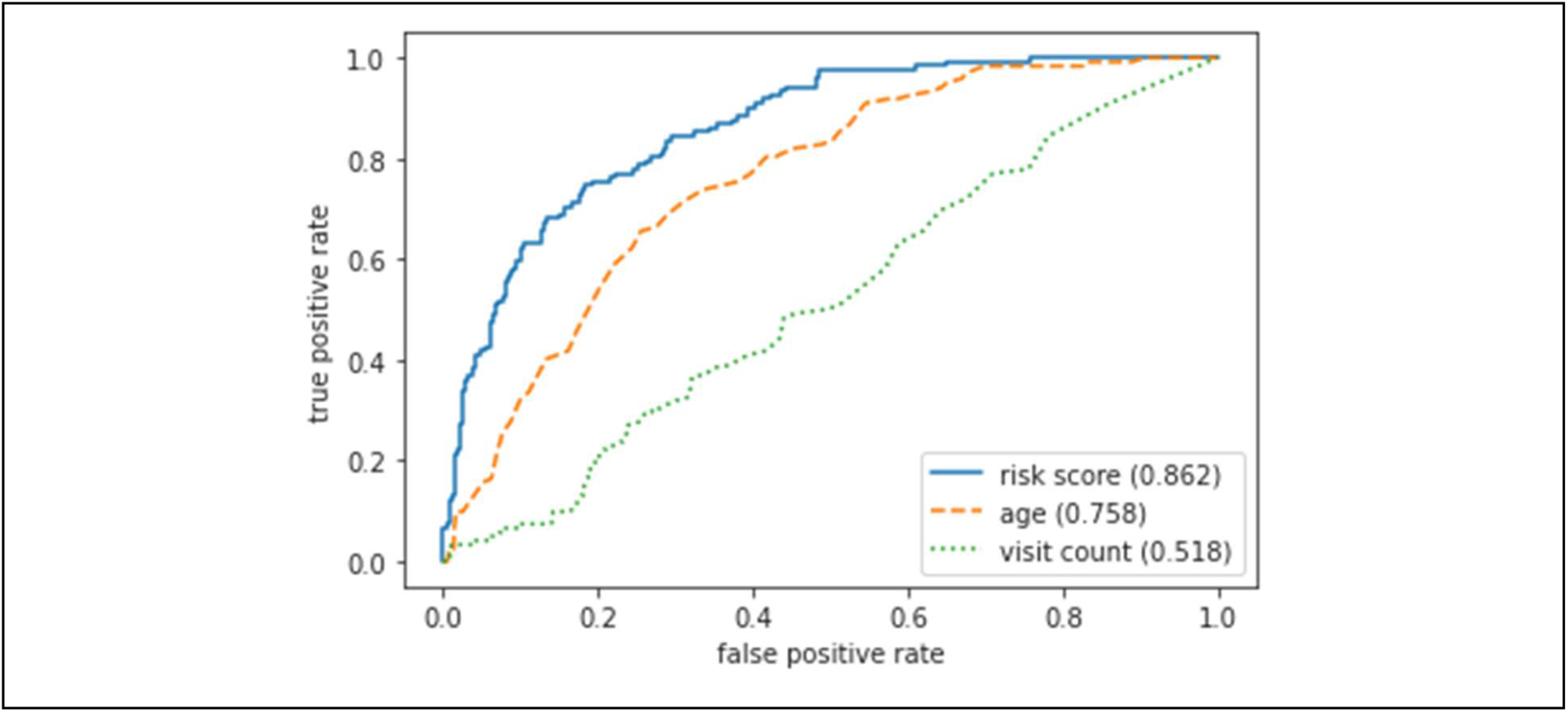
Receiver operating characteristic (ROC) curve of the risk score, age, and historical visit count in predicting the outcome status of the patients. Area under each ROC curve is denoted in the legend.

### Visualization of Patients

The output vector of the fully connected layer in the RNN model is expected to contain information about the patient that is necessary for predicting the risk of developing severe outcomes from COVID-19. We analyzed the patients by visualizing the output vectors of patients on 2-dimensional space using uniform manifold approximation and projection (UMAP)^25^. We trained the RNN model with pre-trained embedding on the entire dataset for 30 epochs and generated output vectors for patients by using the trained model on the entire data. **Figure 5a** shows the scatterplot of the output vectors of all patients in the dataset. **Figure 5b** and **5c** shows the scatterplot of the output vectors of severe patients color labeled by sex (**5b**) and age (**5c**). To further explore the pattern of output vectors for severe patients, we color-labeled them on the 2-dimensional space based on common comorbidities of the cohort. Two common comorbidities of COVID-19 patients in CUIMC, renal failure and type 2 diabetes mellitus (T2DM), were selected^26^. **Figure 6a** and **6b** shows scatterplots of the output vectors of severe patients color-labeled based on the observation of T2DM and renal failure respectively. Scatterplots of the output vectors of male and female severe patients separately color-labeled based on the observation of T2DM and renal failure are shown in **Figure 6c-6f**.

**Figure 5.**
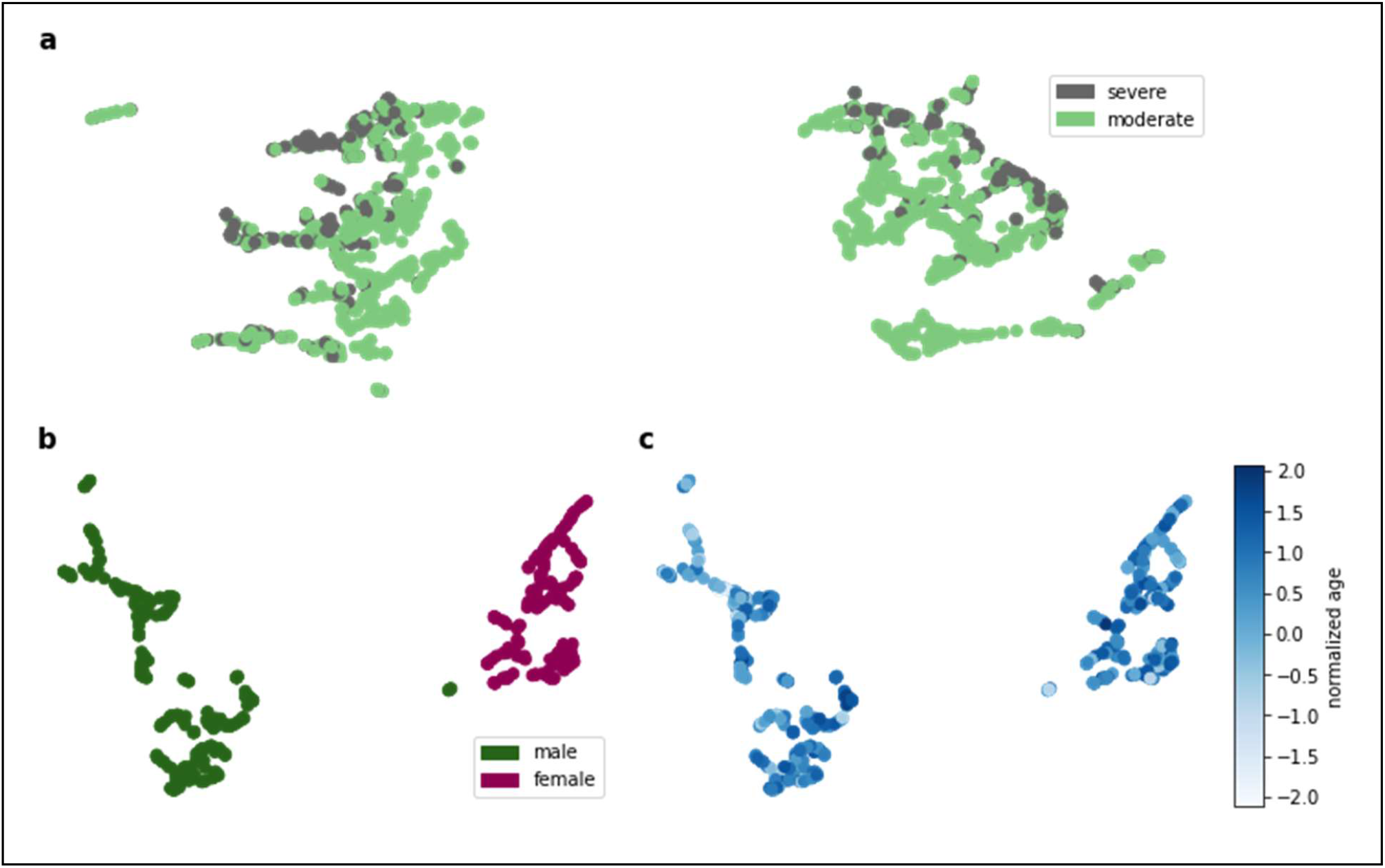
Scatterplots of the output vectors of patients in the COVID-19 cohort. All patients are shown in (**a)** with color representing severity status. Only severe patients are shown in (**b)** and (**c)**, with color representing sex in (**b)** and normalized (i.e. normalized with mean and standard deviation) age in (**c)**.

**Figure 6.**
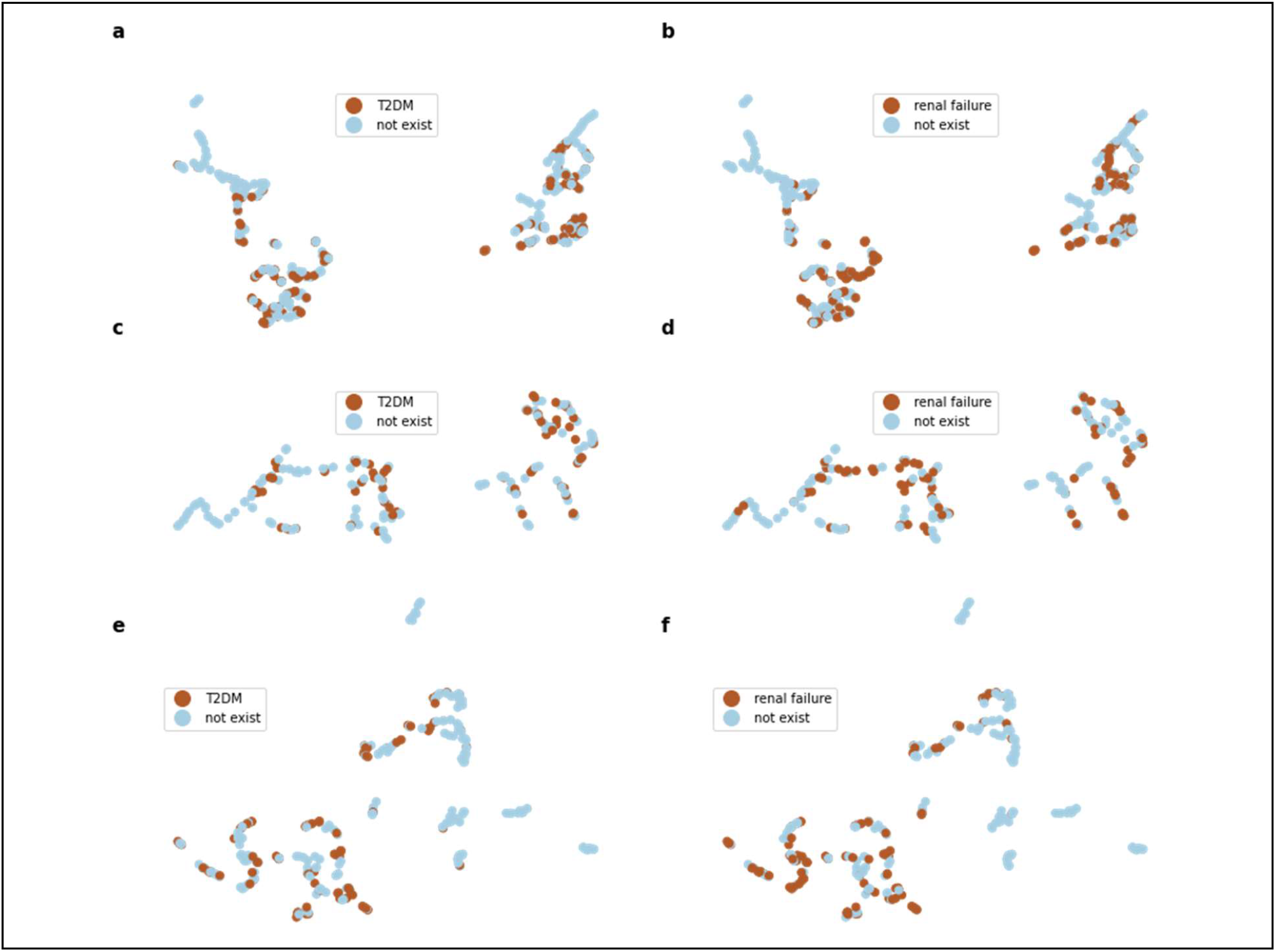
Scatterplots of the output vectors of severe COVID-19 patients, with color representing the observation of (**a**) type 2 diabetes mellitus (T2DM) and (**b**) renal failure. (**c**) and (**d**) are scatterplots of the output vectors of male severe COVID-19 patients, with color representing the observation of T2DM and renal failure respectively. (**e**) and (**f**) are scatterplots of the output vectors of female severe COVID-19 patients, with color representing the observation of T2DM and renal failure respectively.

## DISCUSSIONS

In this study, we proposed an RNN model to predict the risk of developing severe outcomes for COVID-19 patients by utilizing historical EHR data of the patients. The best average AUC was achieved by the RNN model with pre-trained embedding. However, it is worth noting that the difference between average AUC of the RNN model and other baselines are not significant considering the standard error although simple paired t-test confirmed statistically meaningful the difference between the average AUC of the RNN model and other baselines in each initialization scheme. Relatively high standard error is perhaps due to the small size of the dataset. We also found that using randomly initialized embedding in logistic regression and MLP underperforms the models using multi-hot representation as input while using pre-trained embedding improved the performance in all models. This is perhaps because the aggregation across patients’ visits causes information loss for the randomly initialized embedding and the data set was not sufficiently sized to allow the embedding layer to be properly trained starting from random initialization. Additionally, the pre-trained embedding may be suboptimal because we only used the data from the COVID-19 cohort to pre-train the embedding. We expect that the performance will further improve if we use a larger data set to pre-train the embedding.

Although we used a relatively large data set compared to existing COVID-19 studies, which mostly have a few hundred cases^7^, the 2,374 cases in our data set is still considered very small for training deep neural network models that contain a large number of parameters to learn. While the model will be able to learn better with more data, obtaining a large data set, however, is not easy for a single institution due to the limited number of patients (and we certainly hope the number of COVID19 patients will not further increase in our institution). We believe obtaining a larger size of data across different institutions and nations or using other disease cohorts as proxy cohorts will resolve this limitation. One advantage of our approach is that our analysis used a standardized clinical data format, the OMOP Common Data Model. The source code for this analysis can be easily shared with others who have similarly formatted clinical data for evidence aggregation.

While higher accuracies (0.73-0.99) were reported in other studies, the intended use of these models were often not clearly described^7^. The RNN model we propose is intended to aid decision making at the time of or before hospital admission due to COVID-19, since only historical EHR data were needed in the model. In addition, the RNN model can be applied to the general population that is not confirmed COVID-19 positive to identify people at high risk of developing potential severe outcomes if infected by COVID-19. The RNN model can readily be applied to the situations above with much larger datasets or in a real-time setting since it can compute the risk score of the patient in a small amount of time.

We demonstrated the effectiveness of the risk score predicted by the RNN model by analyzing it with basic characteristics (i.e., age and total historical visit count) of the patients. From **Figure 3a,** we can confirm that the risk score is correlated with the patient developing severe outcomes from COVID-19. We found that there exists a statistically significant positive relationship between age and the risk score of the patients in **Figure 3b**, which indicates that age itself is an important factor to predict the outcome status of patients. We also expected that the number of hospital visits in a patient’s medical history would reflect the patient’s general health status and therefore a positive relationship would exist between the total historical visit count and the risk score. **Figure 3c** shows, however, that the relationship between the historical visit count and the risk score of the patients is not strong. The risk score predicted by the RNN model outperforms the other two baselines in predicting outcome status of the patients as shown in **Figure 4**.

From **Figure 5a**, we can see visible clusters of the severe COVID-19 patients. Male and female severe patients were divided into two clusters in **Figure 5b**. Age, however, does not show clearly distinguishable patterns in the clusters from **Figure 5c**. While we cannot confirm clear clusters based on the existence of T2DM or renal failure, we can see that the patients separate into distinct clusters throughout **Figure 6**. Since the patient vectors were generated based on the patients’ observed conditions across visits, these clusters could reflect common comorbidities among severe COVID-19 patients. Additionally, the presence of visible clusters within the scatterplots of the male and female severe patient groups suggests that there exist multiple subgroups of severe COVID-19 patients with distinct characteristics, which shows the potential possibility of subtyping COVID-19 patients. We believe that further efforts to uncover detailed characteristics of the clusters are warranted for subtyping COVID-19 patients.

A drawback of the RNN model is that the model lacks interpretability. The model interpretability is critically important for the model utilizes medical data since interpretable model output can deliver new insights to the problem. For example, we can compare the impact of individual concept on developing severe outcome of COVID-19 by analyzing the weights in logistic regression model with multi-hot vector input. Although the RNN model showed better performance than other models, this gain is at the cost of interpretability. We would like to address this limitation by developing interpretable model without compromising on accuracy in the future study.

Our study shared some common limitations with the existing predictive models for COVID-19 patients. Wynants et al. performed a review of existing predictive models for COVID-19 patients and reported that most of the models have high risk of bias when evaluated with PROBAST (prediction model risk of bias assessment tool)^7,27^. They found that two common causes of risk of bias in predictive models for COVID-19 were lack of external validation and selection bias. Since the COVID-19 cohort in this study includes patients whose clinical course of care has not yet completed and who may still potentially develop a severe outcome, there is a chance that discharged patients without any signal of severe status during hospitalization at NYP/CUIMC will later develop a severe outcome outside of NYP/CUIMC. Future work will include developing an RNN model to predict various states of a patient being infected with COVID-19 rather than simply predicting the risk score. We also plan to modify the RNN model for time-to-event analysis to appropriately handle censored data.

Additionally, the model was not validated with an external cohort. This limitation is mainly caused by medical data exchange issues across different medical institutes, which limits the sharing of medical data across institutions. Since the RNN model is based on a dataset implemented with OMOP common data model, we expect that applying the model to another institution using the common data model will be easily conducted. For example, Burn et al., has performed deep phenotyping on more than 30,000 patients hospitalized with COVID-19 patients in Asian, Europe and American countries using OHDSI network dataset^28^. Future work includes experimenting with and validating the RNN model across different institutions in various countries using the OHDSI network dataset.

## CONCLUSION

We proposed a predictive model using recurrent neural networks to predict the risk of developing severe outcomes for COVID-19 patients. The proposed RNN model outperforms logistic regression and multi-layer perceptron models in predicting severe outcome status of COVID-19 patients. We also demonstrated the effectiveness of the risk score by analyzing the risk score generated by the RNN model with the basic characteristics of the patients. Future work includes experimenting with the model with a larger dataset and validating the model with an external dataset, adding interpretability to the model, as well as further improving the RNN model using more concepts from other domains (e.g., drug, measurements, and procedure) and using time-to-event analysis, which also can address the censored patient issue.

## Data Availability

No data reference.

## ACKNOWLEDGEMENT

This work was supported by The National Library of Medicine grant R01LM012895-03S1 and The National Center for Advancing Translational Science grant 1OT2TR003434-01. Supplementary material is available at https://github.com/Jayaos/rnn-covid. The authors thank the anonymous reviewers for their valuable feedback that helped us to make significant improvements to this study.

