## supplementary material for "Severity Prediction for COVID-19 Patients via Recurrent Neural Networks"

**A. Gated Recurrent Units Preliminaries**

**Figure A1.** depicts the architecture of the Gated Recurrent Units (GRU) cell, where $x_i$, $z_i$, $r_i$, $\tilde{h}_i$, and $h_i$ denote the input, update gate, reset gate, candidate hidden state, and hidden state, respectively, at timestamp $i$. $W_h$, $W_z$, $W_r$, $U_h$, $U_z$, and $U_r$ are the trainable weight matrices. The mathematical formulation of the GRU cell is provided in Eq(1) through Eq(4):

$$z_i = \sigma(W_z x_i + U_z h_{i-1} + b_z) \quad Eq(1)$$
$$r_i = \sigma(W_r x_i + U_r h_{i-1} + b_r) \quad Eq(2)$$
$$\tilde{h}_i = tanh(W_h x_i + r_i \circ U_h h_{i-1} + b_h) \quad Eq(3)$$
$$h_i = z_i \circ h_{i-1} + (1 - z_i) \circ \tilde{h}_i \quad Eq(4)$$

where $\circ$ denotes element-wise multiplication, $tanh()$ denotes the hyperbolic tangent activation function, and $\sigma()$ denotes the sigmoid activation function. The update gate $z_i$ decides how much information should be updated from the input and is computed as Eq(1). Similarly, the reset gate $r_i$ decides how much information should be ignored from the past information and is computed as Eq(2). The candidate hidden state $\tilde{h}_i$ is computed as Eq(3) using the input and the hidden state of the previous timestamp $h_{i-1}$. Finally, the hidden state at timestamp $t_i$ is computed as Eq(4), using the candidate hidden state $\tilde{h}_i$ and the hidden state of the previous timestamp $h_{i-1}$. Since the reset and update gates decide how information from the history of the past inputs should be combined with current inputs to form the new the hidden state, we can utilize the hidden state as the vector containing the information about the patient's total medical history.

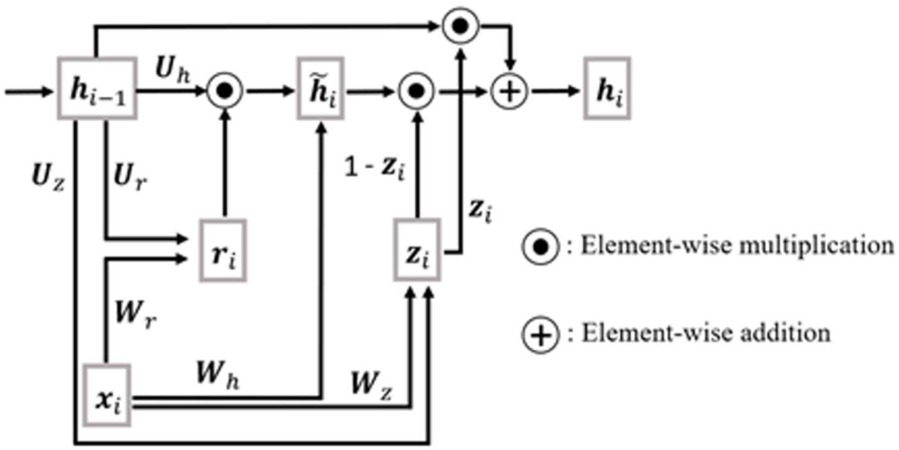


**Figure A1.** The architecture of the Gated Recurrent Units cell.

### B. Hyperparameter Details

**Table B1.** Hyperparameter settings of all models.

| **Model** | **Hyperparameters** |
|---|---|
| Logistic regression w/ multi hot encoded input | Learning rate: 0.00001; $L_2$ weight decay regularization coefficient: 0.001; maximum epochs: 50; batch size: 2 |
| Logistic regression w/ randomly initialized embedding layer | Learning rate: 0.0000005; $L_2$ weight decay regularization coefficient: 0.001; Dimensionality of the embedding: 128; maximum epochs: 50; batch size: 2 |
| Logistic regression w/ pre-trained embedding layer | Learning rate: 0.000005; $L_2$ weight decay regularization coefficient: 0.001; Dimensionality of the embedding: 128; maximum epochs: 50; batch size: 2 |
| Multilayer perceptron w/ multi hot encoded input | Learning rate: 0.000001; $L_2$ weight decay regularization coefficient: 0.001; the number of hidden units of the hidden layer: 128; maximum epochs: 50; batch size: 2 |
| Multilayer perceptron w/ randomly initialized embedding layer | Learning rate: 0.0000005; $L_2$ weight decay regularization coefficient: 0.001; Dimensionality of the embedding: 128; the number of hidden units of the hidden layer: 128; maximum epochs: 50; batch size: 2 |
| Multilayer perceptron w/ pre-trained embedding layer | Learning rate: 0.000001; $L_2$ weight decay regularization coefficient: 0.001; Dimensionality of the embedding: 128; the number of hidden units of the hidden layer: 128; maximum epochs: 50; batch size: 2 |
| Recurrent neural network model w/ randomly initialized embedding layer | Learning rate: 0.00001; $L_2$ weight decay regularization coefficient: 0.001; Dimensionality of the embedding: 128; the number of hidden units of the fully connected layer: 128; maximum epochs: 50; batch size: 2 |
| Recurrent neural network model w/ pre-trained embedding layer | Learning rate: 0.00001; $L_2$ weight decay regularization coefficient: 0.001; Dimensionality of the embedding: 128; the number of hidden units of the fully connected layer: 128; maximum epochs: 50; batch size: 2 |

**C. Training Time**

**Table C1.** Average training time per epoch of all models.

| Model | Training time per epoch (seconds) |
|---|---|
| Logistic regression w/ multi hot encoded input | 5.210 |
| Logistic regression w/ randomly initialized embedding layer | 7.154 |
| Logistic regression w/ pre-trained embedding layer | 7.137 |
| Multilayer perceptron w/ multi hot encoded input | 6.960 |
| Multilayer perceptron w/ randomly initialized embedding layer | 8.610 |
| Multilayer perceptron w/ pre-trained embedding layer | 8.745 |
| Recurrent neural network model w/ randomly initialized embedding layer | 106.118 |
| Recurrent neural network model w/ pre-trained embedding layer | 165.309 |

**D. Values of the average AUC and the standard error in Figure 2**

**Table D1.** Average area under the receiver operating characteristic curve (AUC) and standard error of all models based on 5-fold cross validation.

| Model | Average AUC | Standard error |
|---|---|---|
| Logistic regression w/ multi hot encoded input | 0.783 | 0.041 |
| Logistic regression w/ randomly initialized embedding layer | 0.751 | 0.042 |
| Logistic regression w/ pre-trained embedding layer | 0.801 | 0.030 |
| Multilayer perceptron w/ multi hot encoded input | 0.816 | 0.039 |
| Multilayer perceptron w/ randomly initialized embedding layer | 0.767 | 0.050 |
| Multilayer perceptron w/ pre-trained embedding layer | 0.825 | 0.027 |
| Recurrent neural network model w/ randomly initialized embedding layer | 0.811 | 0.032 |
| Recurrent neural network model w/ pre-trained embedding layer | 0.846 | 0.014 |
